# Single-cell immune survey identifies a novel pathogenic role for T cells in anti-NMDA receptor encephalitis

**DOI:** 10.1101/2024.09.01.24311959

**Authors:** Andrew J. Kwok, Babak Soleimani, Bo Sun, Andrew Fower, Mateusz Makuch, Thomas Johnson, Julian C. Knight, Ho Ko, Belinda Lennox, Sarosh Irani, Lahiru Handunnetthi

## Abstract

We performed single-cell RNA and immune receptor repertoire sequencing of an N-methyl-D-aspartate receptor encephalitis (NMDARE) patient in relapse and remission states, as well as an autoimmune psychosis (AP) patient with anti-NMDAR antibodies. We leveraged publicly available cerebrospinal fluid (CSF) single-cell sequencing data from other neurological disorders to contextualise our findings. Results highlight a key role for T-cells in NMDARE pathogenesis with clonal expansion of both cytotoxic CD4+ and CD8+ effector memory cells in CSF. We further identified interferon responsive B-cells in the CSF during the acute phase of NMDARE and a higher proportion of mononuclear phagocytes in the CSF of AP. Collectively, our work sheds light into the immunobiology of anti-NMDAR antibody-mediated disease.

## Introduction

Patients presenting with N-methyl-D-aspartate receptor antibody encephalitis (NMDARE) develop psychotic symptoms, abnormal movements and seizures. Mechanistically, ovarian teratomas and herpes simplex virus infections may play a central role in disease pathogenesis. Evidence suggests that autoantibodies against the NMDA receptor (NMDAR) are synthesised in ovarian teratomas that act as germinal centres^1^ but how viral infections contribute to the disease remains elusive^2^. Furthermore, little is known about the contribution of immune cells other than B cells to disease pathogenesis.

It has been hypothesised that a subgroup of patients with only psychotic symptoms could represent a *forme fruste* of NMDARE or autoimmune psychosis (AP). This notion is supported by the presence of antibodies against the NMDAR in first episode psychosis^3–5^. Gaining mechanistic insight into the immunopathology of these patients has important clinical implications, as this could lead to the development of effective immunomodulatory treatments.

In this study, we carried out a deep immune cell survey via single-cell RNA (scRNA) and adaptive immune receptor repertoire (AIRR) sequencing of an NMDARE patient during acute relapse and remission states, as well as an AP patient with anti-NMDAR antibodies. We also leveraged publicly available scRNA data from a diverse range of neurological disease controls to contextualise our findings.

## Methods

### Patients and sample processing

We surveyed paired peripheral blood mononuclear cell (PBMC) and cerebrospinal fluid (CSF) compartments of a female in her 30s with NMDARE during an acute relapse and during remission; and a female in her 40s with psychotic symptoms who had serum NMDAR antibodies on two separate occasions (Fig. 1A). All antibody testing was carried out using live cell-based assays as previously described^6^. The patients’ immune cells were loaded onto the Chromium 10X platform for RNA and AIRR library generation, and subsequently sequenced using the NovaSeq6000 platform.

**Figure 1.**
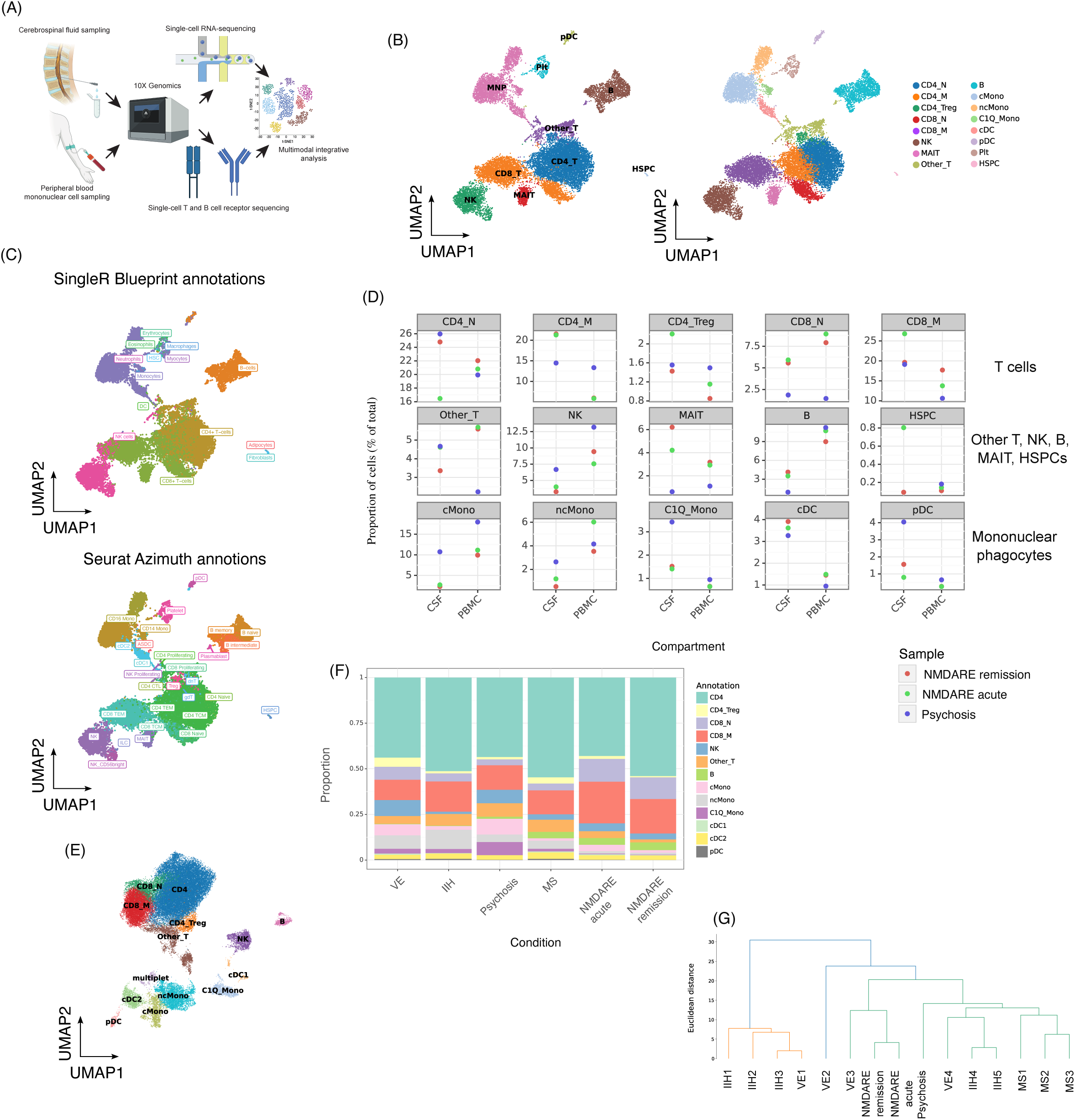
A peripheral blood (PB) and cerebrospinal fluid (CSF) mononuclear cell (MNC) map of NMDARE and autoimmune psychosis (AP). (A) Experimental setup and approach for n = 6 samples (1 NMDARE patient, 1 AP patient). (B) UMAP of broad (left) and fine (right) annotations of 17,110 MNCs from PB and CSF. (C) Algorithmic annotations of MNCs by SingleR (top) and Seurat (bottom) mapping (Methods). (D) Proportions of MNCs in CSF and PB. (E) UMAP of re-integration of 3,342 CSF MNCs with 39,330 cells from publicly available CSF samples of 4 viral encephalitis (VE), 3 multiple sclerosis (MS) and 5 idiopathic intracranial hypertension (IIH). (F) Proportions of MNCs in CSF of NMDARE, AP and publicly available samples. (G) Hierarchical clustering of integrated CSF MNC samples based on cell proportions.

### Data preprocessing and quality control

Gene expression and AIRR data were aligned using CellRanger (v4.0.4). Downstream analysis was performed in scanpy (v.1.9.5). Dimensionality reduction was performed by principal components analysis (PCA). The principal components (PCs) were batch corrected with Harmony (v0.0.9). The top 30 corrected PCs used for downstream clustering and UMAP visualisation.

Expert-driven annotations were performed using curated cell markers and aligned with algorithmic annotations by SingleR (v1.6.1) and Seurat (v4.0.5). Each main cell type from T cells, B cells and mononuclear phagocytes (MNPs) were subsetted out for finer reannotation. Publicly available CSF scRNA-seq data^7, 8^ encompassing 4 viral encephalitis (VE), 3 multiple sclerosis (MS) and 5 idiopathic intracranial hypertension (IIH) samples were reanalyzed with our data with cell and gene QC steps repeated (Supplementary Methods).

### AIRR analysis

AIRR data were analyzed with scirpy (v0.13.1). Clonotype modularity was calculated to identify clonotypes with transcriptomic similarity greater than expected by chance. TCR clonotypes were analyzed for antigen binding by querying the vdjdb database. Differential gene expression analysis was performed by selecting the clonotypes with the highest modularity scores and comparing the gene expression of these cells with relevant groups i.e. memory CD8+ T cells (Supplementary Methods).

### Interferon responsive gene set enrichment

Gene sets representing responsiveness to interferon (IFN) stimulation and signalling were collated^9–12^. Patient derived gene expression profiles were scored for enrichment of IFN responsive gene sets using the scanpy.tl.score_genes function.

## Results

### Proportional differences in immune cell populations are observed across disease states

We sequenced a total of 17,110 cells and recapitulated key mononuclear cell (MNC) types as shown in Figure 1. There was concordance between our manual annotations and results from data-driven approaches (Fig. 1B and 1C).

We noted several clear proportional differences in cell populations across disease states. First, in both NMDARE acute and remission states, CD4+ memory T cell (CD4_M) proportions were elevated in CSF and correspondingly lower in PBMCs (Fig. 1D). By contrast, CD4+ memory T cell proportions were similar in both CSF and PBMCs in AP (Fig. 1D). Second, there were more CD8+ memory (CD8_M) cells in CSF and correspondingly fewer in PBMCs in acute compared to remission NMDARE (Fig. 1D). Third, we found a higher proportion of MNPs including classical monocytes (cMono), C1Q-high monocytes (C1Q mono) and plasmacytoid dendritic cells (pDCs) in CSF of the AP patient than in either acute or remission states of the NMDARE (Fig 1D). After integrating our CSF cells with publicly available CSF scRNA-seq data (Fig. 1E, Supplementary Methods), overall patterns and hierarchical clustering of cell type proportions did not show any clear similarities between the NMDARE and AP samples (Fig. 1F and G). Both NMDRE and AP had MNC profiles that differed from autoimmune driven MS and non-immune IIH (Fig. 1F and G).

### Memory CD8+ T cells, activated CD4+ T cells and cross presenting dendritic cells are elevated in acute NMDARE CSF

We subsetted each cell lineage for finer resolution analysis (Fig. 2 and Supplementary Fig. 3-4) and discovered two effector CD4+ populations expressing *CXCR3* (i) CD4_CXCR3+_TBX1+, and (ii) CD4 CTL proliferation (CD4_CTL_prolif) (Fig. 2B and 2C). The CD4 CTL proliferation cluster exhibited granzyme gene expression and high *PDCD1* and *CTLA4* levels, suggesting a particularly cytotoxic and activated phenotype (Fig. 2C). These CD4+ T cells were expanded in the CSF compartment of both NMDARE states compared to the AP patient (Fig. 2D). Similarly, in the CD8+ compartment, effector memory cells (CD8_EM) with high expression of terminal effector function associated granzyme genes (*GZMB*, *GZMH*) and perforin (*PRF1*) (Fig. 2C) were higher in CSF of the acute NMDARE state than in remission (Fig. 2D). Expansion of both effector CD4+ and effector memory CD8+ T cells was the greatest in NMDARE compared to other neurological conditions, including MS (Supplementary Fig. 5, Fig. 2E). Commensurate with the CD8+ memory T expansion, we identified increased cross-presenting of conventional dendritic cells (cDCs; cDC_XCR1+) in NMDARE CSF, particularly in the acute state (Supplementary Fig. 4C).

**Figure 2.**
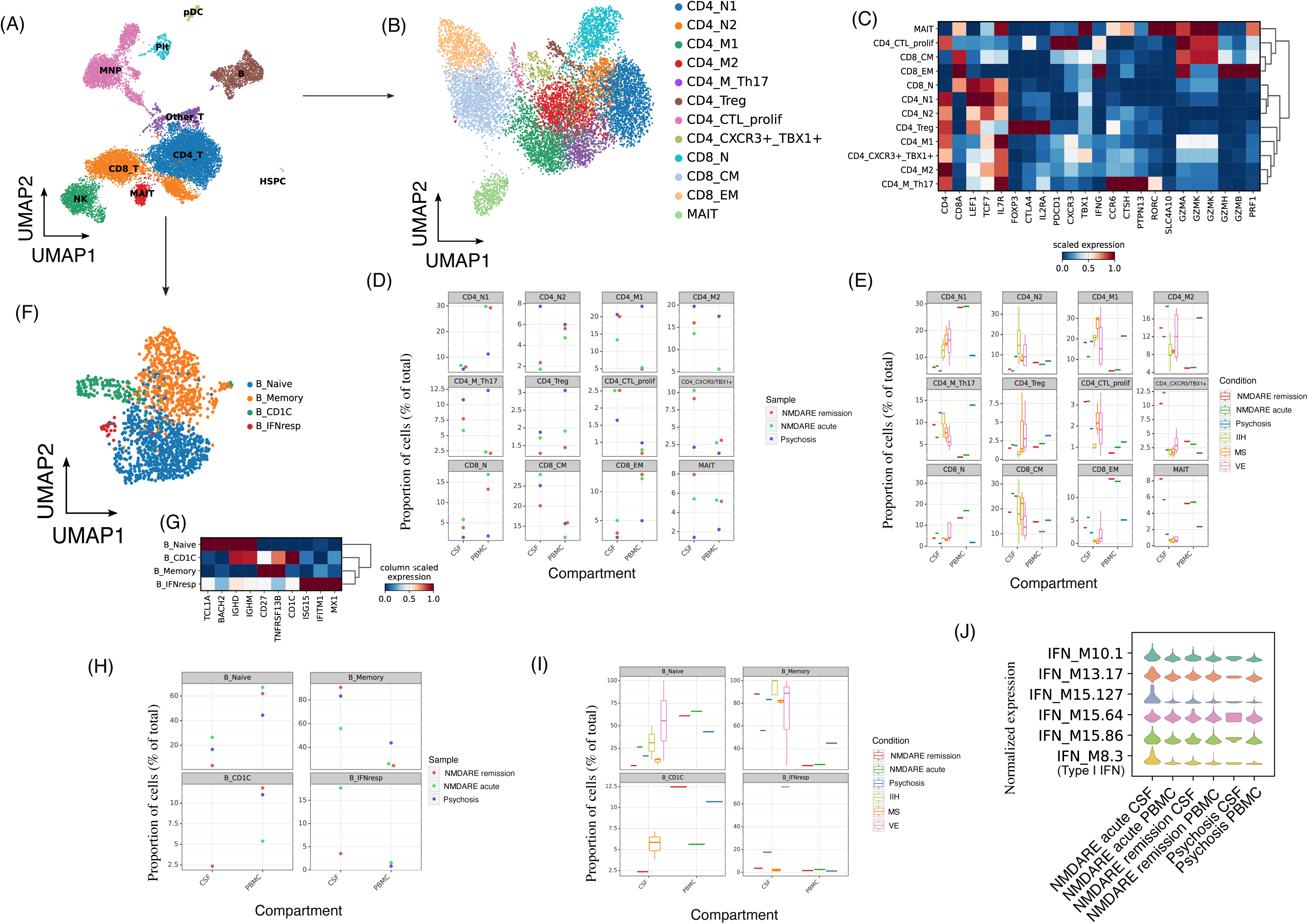
Fine-resolution analysis of adaptive immune cells (T and B cells). (A) Subsetting, re-processing and reclustering of T and B cells. (B) UMAP of peripheral blood (PB) and cerebrospinal fluid (CSF) T cells after reclustering. (C) Marker gene heatmap for T cell subsets. (D) Proportions of T cells in CSF and PB of NMDARE and AP. (E) Proportions of T cells from NMDARE and AP samples as compared to CSF samples from viral encephalitis (VE), multiple sclerosis (MS) and idiopathic intracranial hypertension (IIH). (F-I) As per B-E, for B cells. (J) Normalised expression of six whole-blood transcriptome-derived interferon gene modules^11, 12^ per NMDARE / AP sample.

### Appearance of IFN-responsive B cells in NMDARE CSF

Subsetting of B cells revealed a cluster that exhibited high expression of IFN-stimulated genes (*MX1*, *IFITM1*, *ISG15*) (Fig. 2F and 2G), which we annotated as IFN-responsive B cells (B IFNresp). Interestingly, B IFNresp cells were more abundant in CSF than in the periphery, and substantially more so in the acute vs. remission phase of NMDARE (Fig. 2H). There were no B IFNresp cells in the CSF compartment of the AP patient (Fig. 2H) but similar cells could be found in a viral encephalitis patient at a particularly high proportion (Supplementary Fig. 5, Fig. 2I). To validate the IFN responsiveness of these B cells, we scored the B cells for enrichment of published IFN-responsive gene sets^9, 10^. We found that the B IFNresp cells indeed showed a higher expression of these genes than all other subsets of B cells (Supplementary Fig. 6, Fig. 2J).

### NMDARE CD4+ and CD8+ T cells exhibit clonal expansion and share receptor reactivity with viral antigens

In the NMDARE samples, we detected clonal expansion in both CD8+ and CD4+ T cells (Fig. 3A). In absolute numbers, we noted the greatest clonal expansion in CD8+ central memory (CD8_CM) cells, while as a fraction of the cell subset, CD8+ EM cells displayed the greatest expansion (Fig. 3B). The effector-like CD4+ subset, the CD4 CTL proliferating cells, was the most clonally expanded within the CD4+ compartment (Fig. 3B).

**Figure 3.**
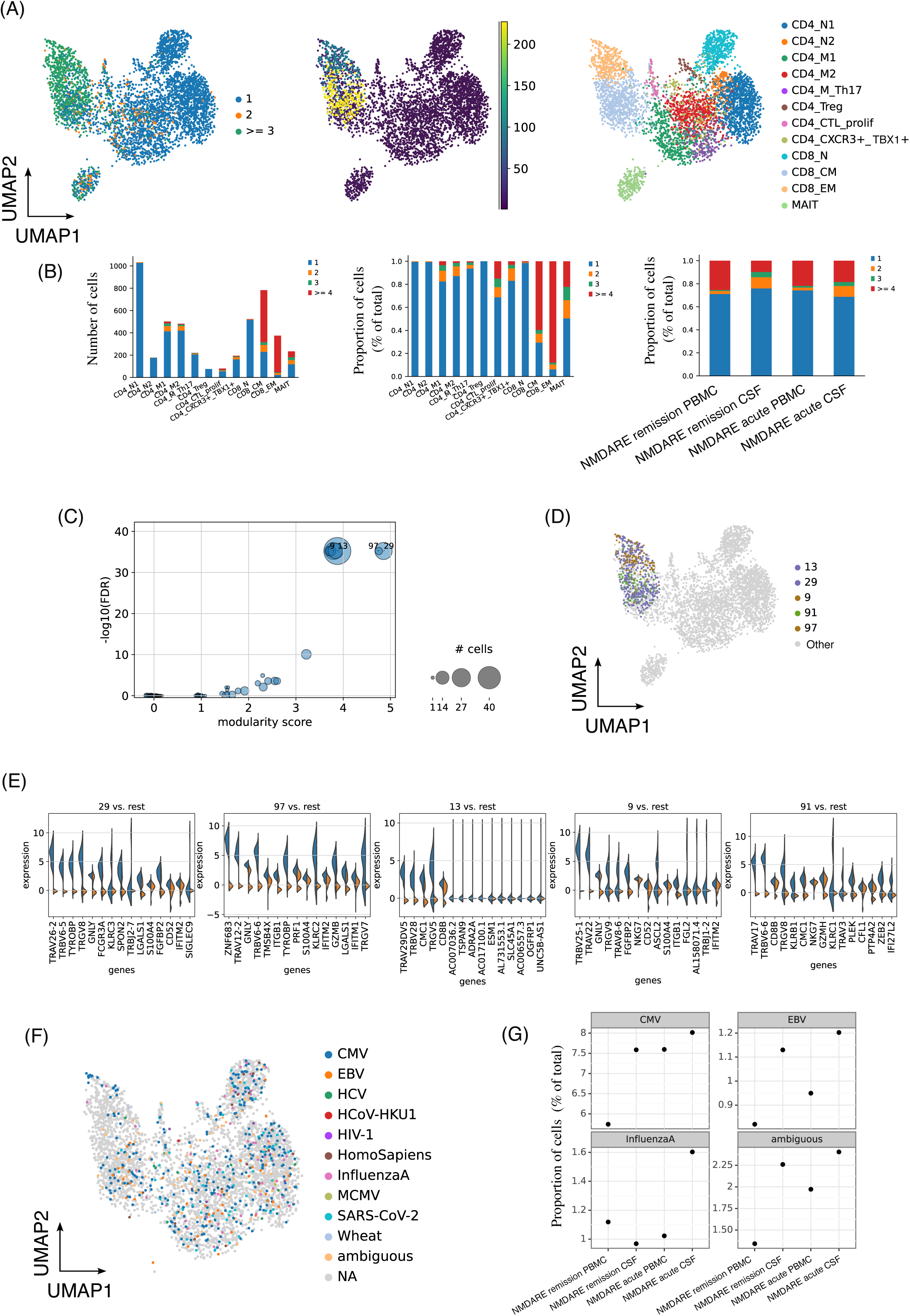
T cell receptor and clonal expansion analysis in NMDARE. (A) UMAPs of T cells colored by T cell clonal expansion by clone ID size (left and middle) and T cell subset annotation (right). (B) Clonally expanded T cells in absolute numbers (left), as a total proportion of each subset (middle) and as a total proportion of each sample (right). (C) Modularity score analysis of T cell clones (Supplementary Methods). (D) Top 5 clonotypes of highest modularity on UMAP. (E) Top differentially expressed genes between top 5 clones of highest modularity vs. other CD8 memory T cells. (F) Antigen species recognised by each T cell clone. (G) Proportions of antigens recognised by T cells (at least 1%) in each sample.

To test whether any of these expanded T cell clones exhibited similarities in gene expression profiles, we calculated modularity scores (Fig. 3C, Supplementary Methods). The five top clonotypes by modularity scores and statistical significance arose from the CD8+ compartment (Fig. 3D). The clones of high modularity showed an increase in granzyme genes (*GNLY*, *GZMB*, *GZMH*), suggesting heightened cytotoxic effector function (Fig. 3E). Two clonotypes (97 and 9) also showed a greater expression of *ITGB1*, a marker of CNS homing cells (Fig. 3E).

Finally, we assessed antigen specificity of T-cell clonotypes in NMADRE using a curated repository (Fig 3F). We observed several T cells from NMDARE samples exhibited binding against herpes viruses, namely CMV and EBV epitopes (Fig. 3G). T cells from NMDARE in the acute phase also exhibited an overlap with Influenza A epitopes (Fig. 3G).

## Discussion

Here, we present a single-cell transcriptomic and immune repertoire atlas of peripheral blood and CSF MNCs from patients with anti-NMDAR antibody-mediated pathology. Importantly, we found that immune profiles differed both between the NMDARE and AP patients, as well as between relapse and remission states of NMDARE. Furthermore, neither NMDRE nor AP immune profiles resembled classic CNS autoimmune disorders such as MS or non-immune conditions such as IIH.

We highlight a novel role for T cells in NMDARE pathogenesis. Specifically, we noted clonal expansion in both CD8+ and CD4+ T cells. The CD8+ effector memory cells displayed the greatest expansion with heightened cytotoxic and chemotaxic characteristics during acute relapse. Further, the antigen specificity of NMDARE T-cell clonotypes highlights a potential mechanism through which viral infections could contribute to disease mechanisms. Another key finding was the appearance of IFN-responsive B cells in the CSF compartment particularly during the acute phase of NMDARE. It is possible that type 1 IFNs could contribute to the disease through facilitating B-cell proliferation and differentiation^13^.

We acknowledge that our study has limitations. This study had a low number of samples and larger studies are needed to confirm the generalisability of our findings. Nevertheless, we integrated our data with a larger number of samples from publicly available datasets to improve the robustness of our analysis. Additionally, our results are observational in nature and future functional experiments, e.g. assays to confirm the epitope binding of clonally expanded T cells in NMDARE patients, will be needed.

In conclusion, we present a small-scale but valuable reference of peripheral blood and CSF MNCs in NMDARE and an AP patient, highlighting the relevance of T cells and viruses in NMDARE disease pathogenesis. These findings carry implications for treatment strategies in NMDARE.

## Data Availability

Raw and processed data and code used for analysis will be made available upon acceptance of the manuscript.

## Acknowledgements

We thank the patients who participated for their generous contribution to this study, and the clinical staff, and Oxford University NHS Hospital Trusts recruiting hospitals involved in patient recruitment and sample collection. LH is supported by the National Institute for Health and Care Research (NIHR) Oxford Health Biomedical Research Centre United Kingdom, and a pump priming grant from the University of Oxford. SRI is supported by a senior clinical fellowship from the Medical Research Council [MR/V007173/1].

## Author contributions

Conceptualization: LH; Data curation: AJK; Formal analysis: AJK, LH; Funding acquisition: LH; Investigation: AJK, LH, Methodology: AJK, LH, BSo, BSu, AF, MM, TJ; Project administration: AJK, LH; Resources: AJK, JCK, HK, BL, SI, LH; Software: AJK; Supervision: SI, JCK, LH; Validation: AJK, LH; Visualization: AJK, LH; Writing – original draft: AJK, LH; Writing – review & editing: AJK, LH, BSo, BSu, AF, MM, TJ, JCK, HK, BL, SI

---

**Supplementary Figure 1.**
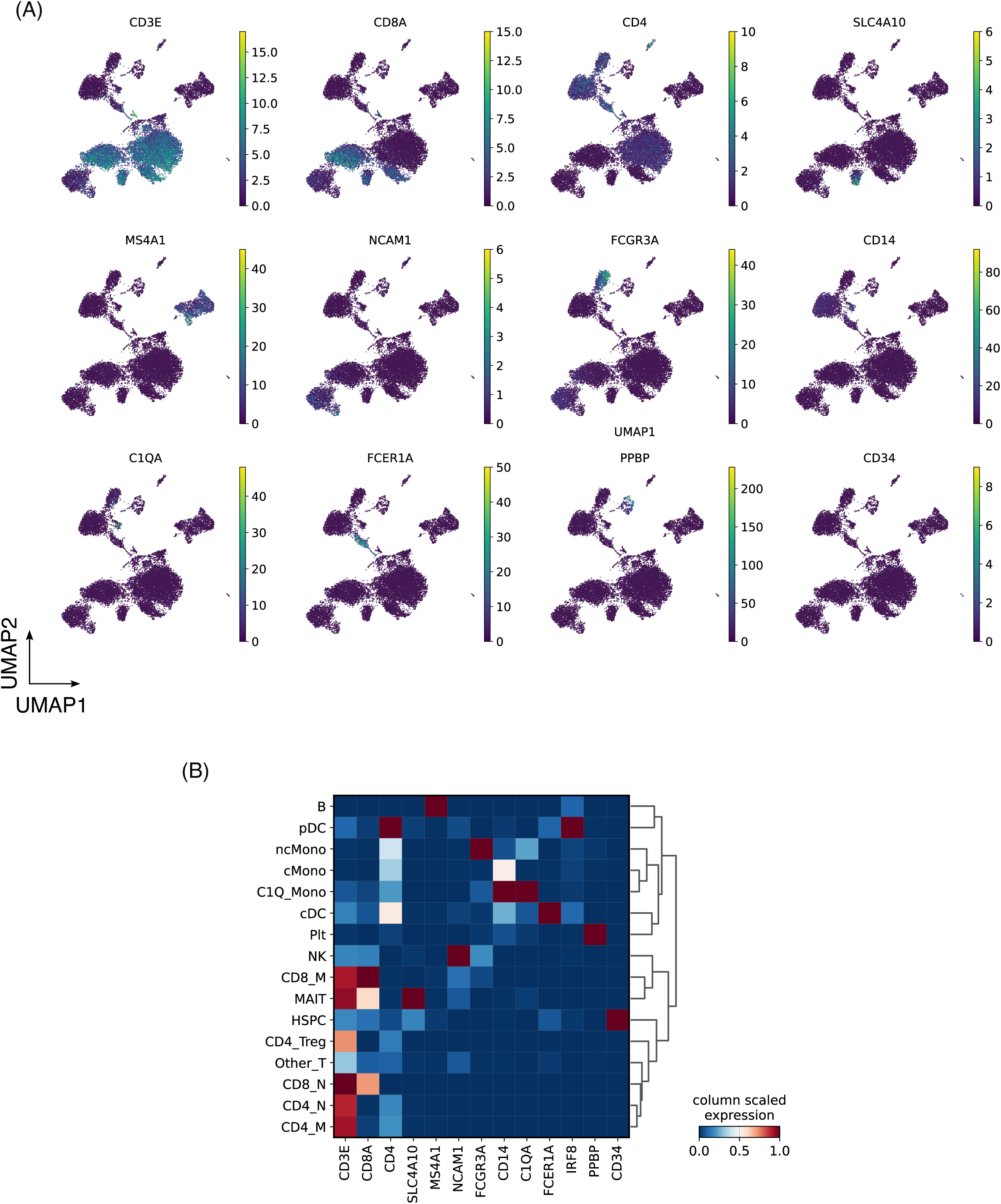
NMDARE / AP MNC markers. (A) UMAPs of 17,110 peripheral blood and cerebrospinal fluid mononuclear cells from n = 6 samples (1 NMDARE patient, 1 AP patient) with normalised cell lineage marker gene expression overlaid. (B) Heatmap of lineage marker gene expression per broad annotated cell population.

**Supplementary Figure 2.**
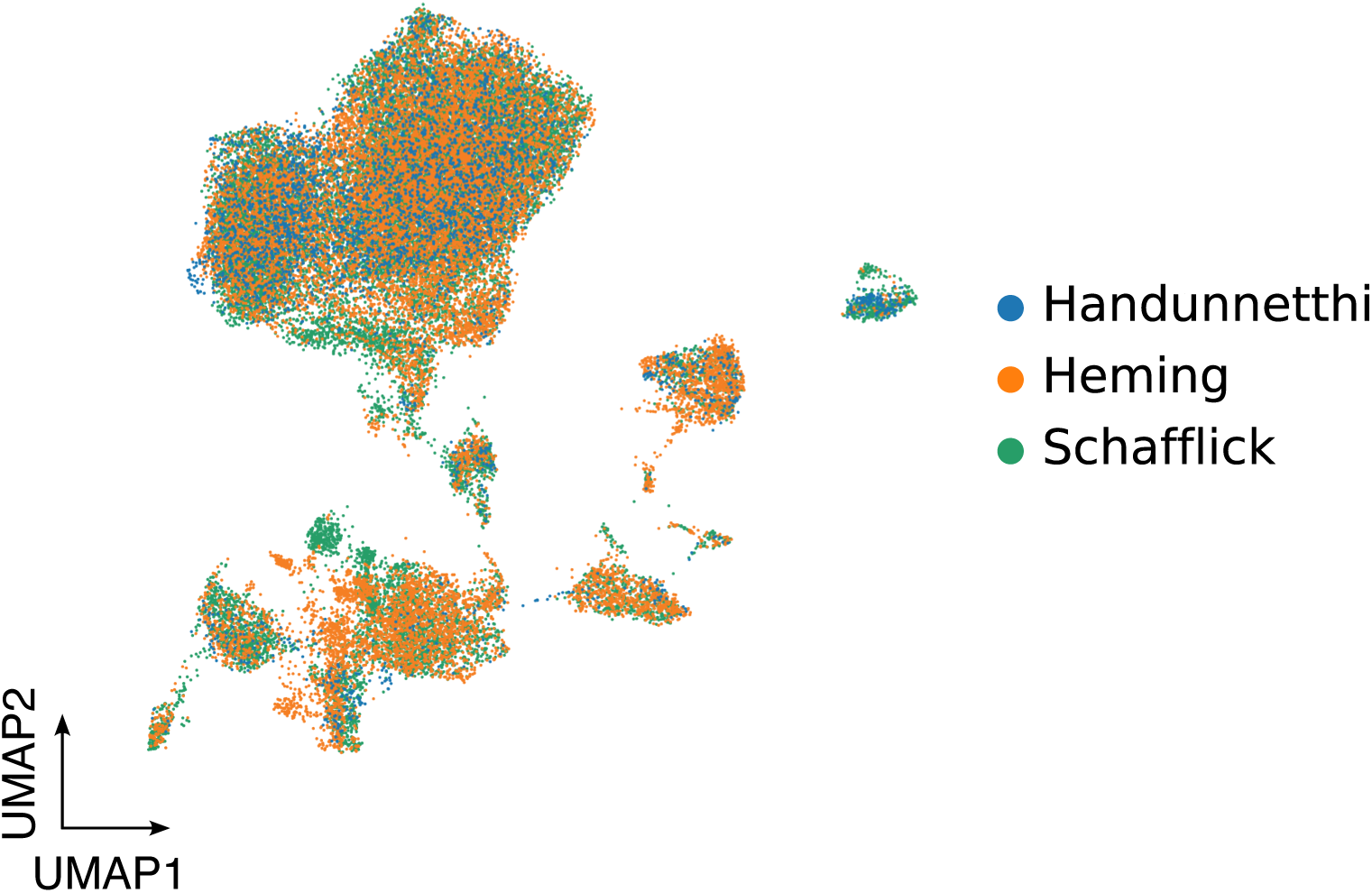
CSF cell integration. UMAP of 33,672 CSF MNCs integrated together (3,342 from current dataset, plus 39,330 publicly available) colored per batch/dataset.

**Supplementary Figure 3.**
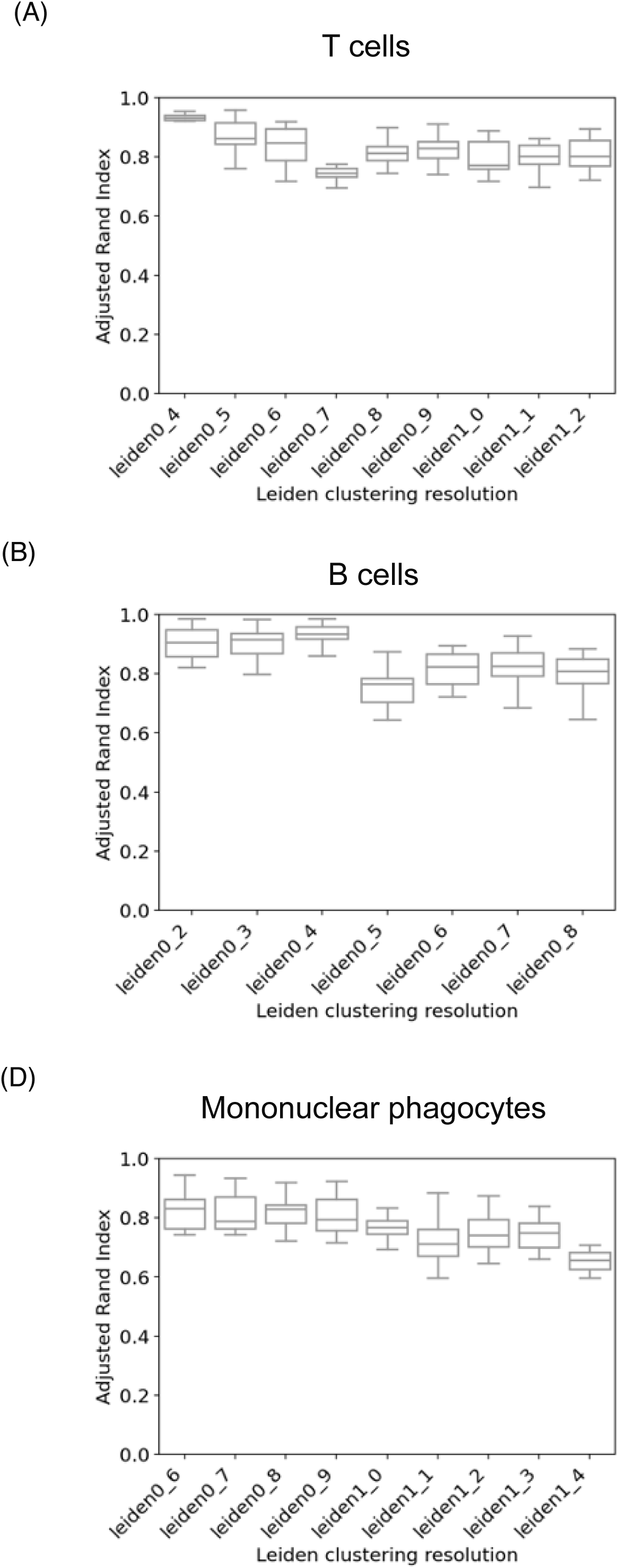
Subclustering resolution analysis (Supplementary Methods). The adjusted rand index (ARI) was plotted against each clustering resolution to the select highest clustering resolution at which ARI begins to drop off for (A) T cells (B) B cells and (C) mononuclear phagocytes.

**Supplementary Figure 4.**
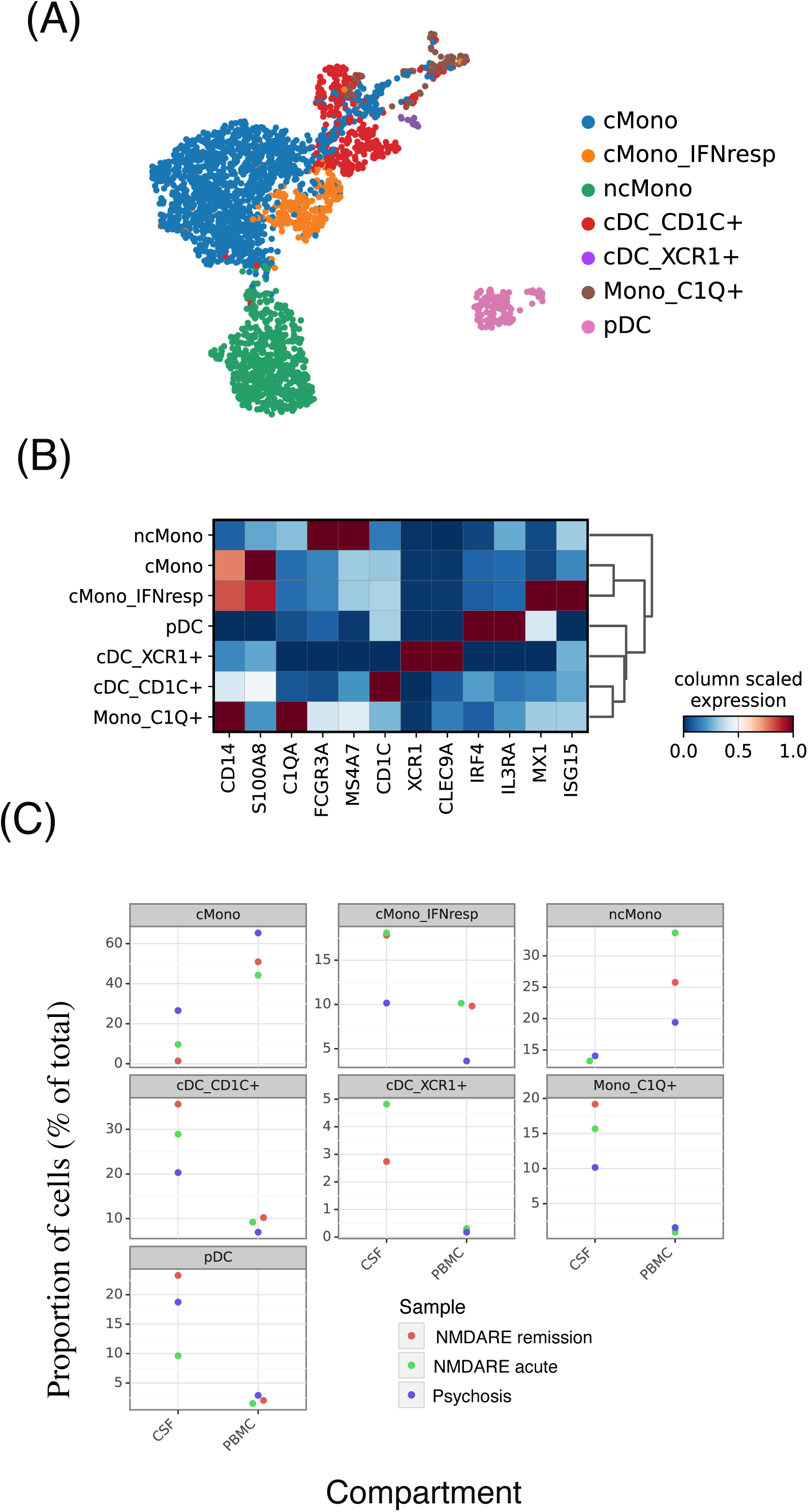
Fine resolution analysis of mononuclear phagocytes (MNPs). (A) UMAP of MNPs after reclustering. (B) Marker gene heatmap for MNP subsets. (C) Proportions of MNPs in CSF and PB of NMDARE and AP.

**Supplementary Figure 5.**
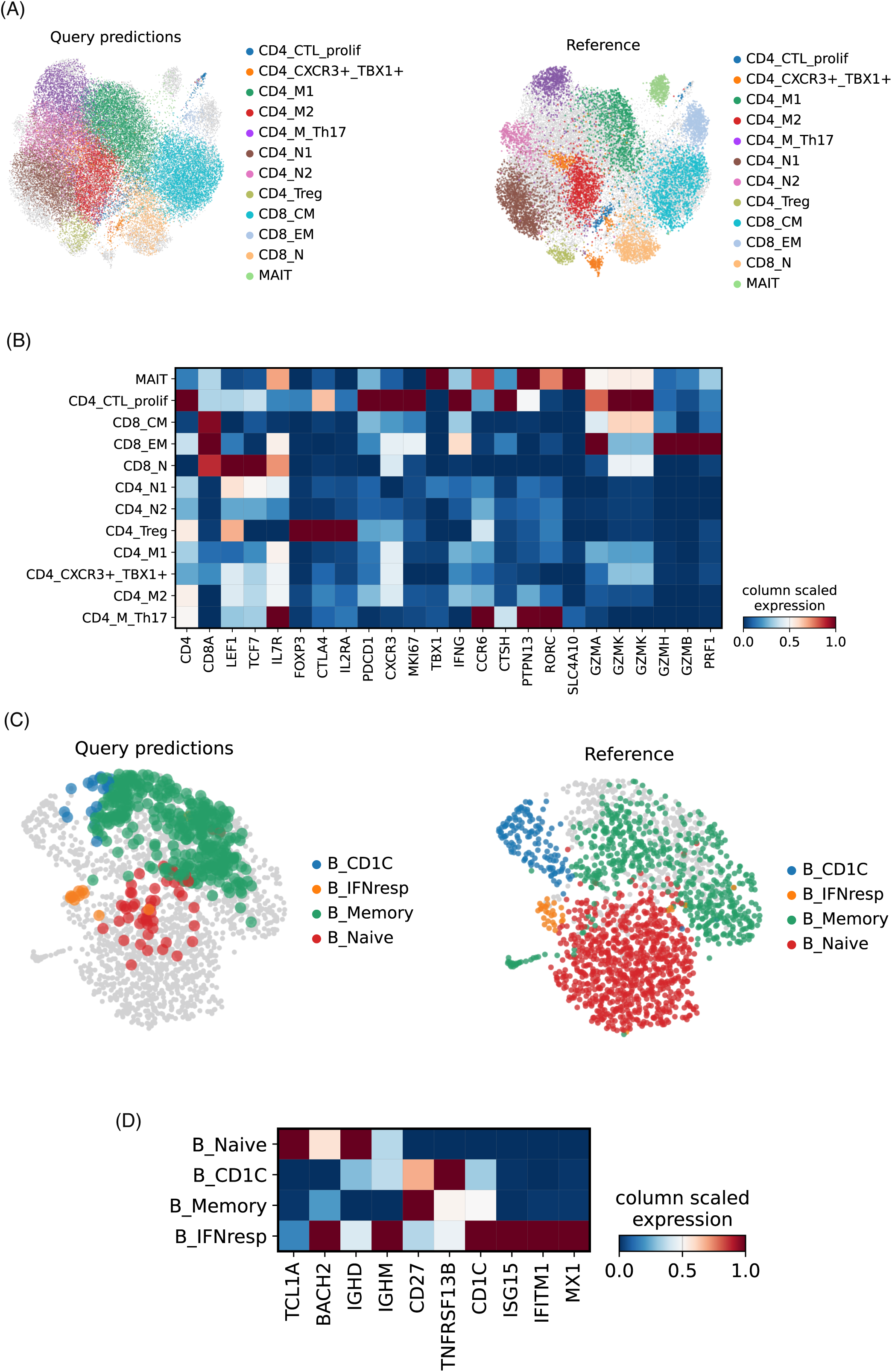
CSF adaptive immune cell reference mapping. (A) UMAPs after T cells from the publicly available CSF datasets (query, left) were mapped to NMDARE and AP T cells as a reference (right). (B) Marker gene heatmap for query T cell clusters. (C-D) As per A-B, for B cells.

**Supplementary Figure 6.**
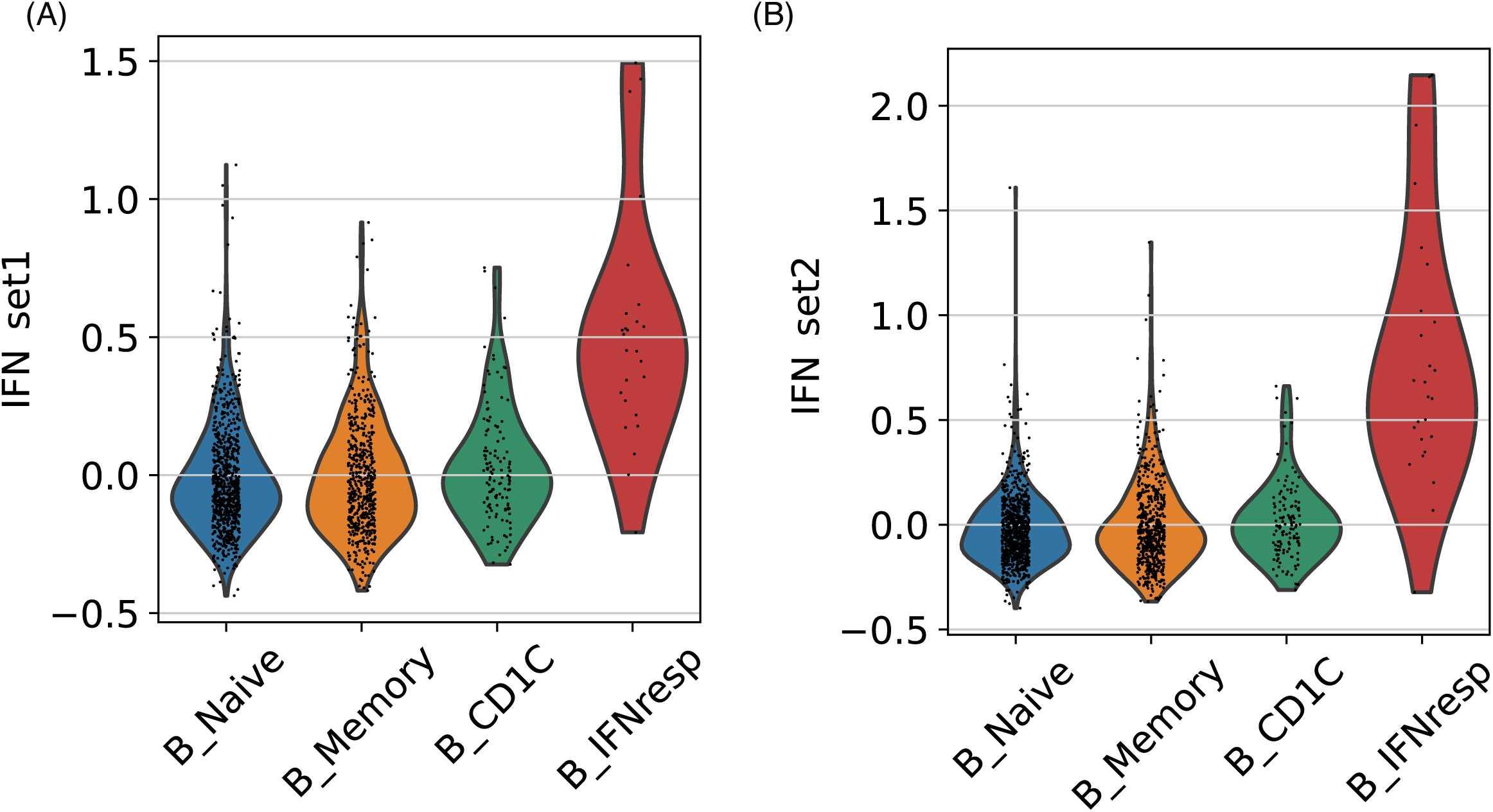
Interferon (IFN) gene set analysis. Normalised expression of two type I IFN responsive gene sets per B cell subset (A^10^, B^9^).

