## Supplementary_methods for "Single-cell immune survey identifies a novel pathogenic role for T cells in anti-NMDA receptor encephalitis"

*Patients and sample processing*

Patients were recruited from Oxford University Hospitals into a study approved by the Yorkshire/Humber Research Ethics Committee in the United Kingdom (REC16/YH/0013). These patients were not taking any immunotherapy at the time of sample collection. Paired CSF and BMBC samples were collected and processed immediately for storage in liquid nitrogen. Later, cells were thawed, resuspended and loaded onto the Chromium 10X platform. Sequencing was performed on the NovaSeq6000 platform with the 100-cycle NovaSeq v1.5 reagent kit.

*Data preprocessing and quality control*

Cells expressing <100 or >4,000 genes, >20% mitochondrial reads, or >2% hemoglobin reads were removed. Genes expressed in <10 cells or with a total count <3 were removed. Scrublet (v.0.2.3) and doublet detect (v.3.0) were both used on default settings to remove doublets with automatic thresholds. Counts were normalized to 10,000 reads per cell and then log normalised. Excluding T cell receptor, B cell receptor and HLA genes, 3,000 highly variable genes (HVGs), were identified using the Seurat vst algorithm (scanpy v.1.9.5).

*Clustering and cell annotations*

All PBMCs and CSF cells were clustered and annotated together initially. Expert driven annotations were supplemented by literature curated cell type markers.

Annotations in SingleR utilized both Human Primary Cell Atlas and Blueprint reference cell types. Seurat annotations were performed by PCA mapping against the reference Azimuth PBMC dataset with default settings.

In the further rounds of preprocessing, subclustering and finer reannotation for each lineage, subsetted cells were first reprocessed, clustered and annotated, with contaminant populations and multiplets not identified in initial QC discarded before a third round of reprocessing and clustering was performed. Multiplets were defined here as clusters which expressed markers of more than one lineage.

New UMAP embeddings were recalculated but cells were not reannotated in this third round - we reasoned that if the new UMAP embeddings continued to show separation of the cells by the clusters originally assigned in the second round pre-contaminant/multiplet removal, then this indicated the annotations and clustering in the second round were biologically valid and robust. Reannotations were done through expert immunological knowledge driven analysis.

To decide on the resolution for clustering, the data were subsampled without replacement and re-clustered (90% of cells, 20 iterations). The adjusted Rand index (ARI) for the original vs. the subsampled clustering outcomes were computed. The resolution just before the ARI began to drop (indicating a decrease in robustness) was used for downstream analysis (Supplementary Fig. 3).

*Integrative CSF analysis*

We downloaded publicly available CSF scRNA-seq data from Schafflick et al[1] and Heming et al[2]. Post cell calling files were used for analysis. Cells from these datasets were reanalysed with our data with cell and gene QC steps repeated. We found that one viral encephalitis, four IIH, and seven MS samples integrated poorly based on both clustering and UMAP visualisation, especially in terms of T cells. We therefore filtered these samples out and reprocessed and reclustered the data. 50 PCs were Harmony batch corrected and taken for downstream clustering and UMAP visualisation. The 20 clusters were merged and annotated for identification of the main cell types. We further filtered out neuro-COVID samples for downstream analyses.

Aggolmerative hierarchical clustering of the samples was performed on the cell proportion matrix. The cell proportions were first quantile normalised per sample, before being clustered with a Euclidean distance metric and Ward's linkage.

*Publicly available CSF data reference mapping*

To align the publicly available CSF scRNA-seq data from Schafflick et al[1] and Heming et al[2] against our per lineage fine annotations, we employed a reference mapping approach. We employed the scArches method as applied to scANVI (single-cell annotation using variational inference) to map adaptive immune (T and B) cells from the publicly available CSF data (query cells) to our subsetted and fine annotated T and B cells (reference cells), with UMAP visualisations and marker gene heatmaps confirming accuracy of the reference mapping. Query T and B cells were selected based on the annotations of the integrative CSF analysis.

*Adaptive immune receptor repertoire analysis*

QC was performed with the default scirpy immune receptor model where single or dual receptor cells were retained and multi-chain cells were discarded. Clonotypes were defined as cells with identical CDR3 amino acid sequences and were called publicly.

Clonotype modularity was calculated using the scirpy.tl.clonotype_modularity function. Cells with a high clonotype modularity are transcriptionally more similar to each other than by random. In the differential gene expression analysis, the choice of not comparing the CD8+ clonotypes with the highest modularity scores to naive CD8+ T cells was made to ensure that differentially expressed genes identified would not simply reflect a naive-memory difference.
